# Ferric carboxymaltose has a higher distribution into myocardium than gadobutrol – a quantitative T1 mapping study in healthy volunteers

**DOI:** 10.1101/2023.03.01.23285660

**Authors:** Goran Abdula, Magnus Lundin, Jannike Nickander, Peder Sörensson, Andreas Sigfridsson, Raquel Themudo, Martin Ugander

## Abstract

**Objectives:** The dynamic tissue distribution of the clinically available intravenous iron substitution agent ferric carboxymaltose (FCM) is largely unknown. We aimed to evaluate and quantify the dynamic tissue distribution of FCM using cardiovascular magnetic resonance (CMR).

**Materials and methods:** T1 mapping was prospectively performed to determine T1 and the partition coefficient (lambda) in myocardium, liver, spleen and skeletal muscle up to 60 minutes after onset of a 15-minute-long infusion of 20 ml (50 mg iron/ml) FCM in healthy male volunteers. For comparison, myocardial lambda for gadobutrol (0.2 mmol/kg) was measured in a separate group of age-matched healthy male volunteers. The t-test was used for group comparisons.

**Results:** A total of 25 healthy males (mean±SD age 27±3 years) underwent CMR with intravenous FCM (n=8) or gadobutrol (n=17). T1 values decreased in myocardium, blood, liver, spleen and skeletal muscle following FCM infusion (*p* < 0.001 for all). Lambda for FCM in myocardium and spleen remained constant over time after injection of FCM (mean±SEM 64±8% and 81±20%, respectively, at 30 minutes), while liver and skeletal muscle lambda increased. Myocardial lambda for FCM was higher than myocardial lambda for gadobutrol (64±8 vs 45±1%, p=0.003).

**Conclusions:** T1 mapping can detect and quantify the dynamic tissue distribution of iron from FCM in the myocardium, liver, spleen, and skeletal muscle. Higher myocardial lambda for FCM compared to gadobutrol, indicating that FCM distributes to a greater extent into the myocardium than extracellular contrast agents, most likely due to additional distribution into the intracellular space.

**Key points:**

- Understanding iron physiology in heart failure is crucial, as intravenous iron therapy by ferric carboxymaltose (FCM) improves cardiac function.
- T1 mapping effectively measures FCM in myocardium, showing higher myocardial distribution than gadobutrol, likely due to additional intracellular distribution.
- This technique offers new insights into myocardial iron physiology in both healthy and diseased hearts.

## Introduction

Iron deficiency is a common comorbidity in patients with heart failure [1]. FCM is an intravenous iron preparation which enables controlled release of non-ionic iron within the cells into the serum under physiological condition [2]. Studies reveal that treatment with intravenous FCM in patients with heart failure and iron deficiency improves clinical symptoms and functional capacity, and decrease hospital admission regardless of the presence of anemia [3; 4]. This observation suggests that iron deficiency has a pathophysiological mechanism beyond anemia that contributes to heart failure. Recently, myocardial iron deficiency has been suggested to have a role in the development of heart failure [5]. Therefore, the ability to non-invasively quantify myocardial iron uptake is of considerable clinical interest, since the pathological consequences of iron deficiency are potentially reversible.

The current non-invasive reference method for quantification of myocardial iron relies on using magnetic resonance imaging to measure T2*. The normal myocardial T2* is typically 37±5 ms. However, a threshold of 20 ms, corresponding to double the amount of myocardial iron than normal [6], has been used because T2* at low iron levels is sensitive to non-iron influences, such as motion and susceptibility artefacts [7]. Recently it has been demonstrated that T1 mapping can be used to quantify iron concentrations [8]. In addition, T1 mapping has the potential to detect milder degrees of myocardial iron overload compared to T2* [9]. Approximately one third of subjects with low myocardial T1, secondary to iron overload in the myocardium, had normal T2* values (>20 ms) [9]. Furthermore, myocardial T1 mapping methods are less prone to image artifacts compared to T2* sequences [10]. Post-contrast T1 mapping has been used to quantify gadolinium-based contrast agent concentration over time [11; 12], and its distribution volume in the myocardium [13]. Pre- and postcontrast T1 measurements have been used to calculate the blood-tissue partition coefficient, lambda (λ), for gadolinium-based contrast agents [14]. However, T1 mapping has not been used to quantify changes in tissue iron content in vivo after intravenous administration of iron.

Therefore, the purpose of this study was to evaluate the use of T1 mapping to detect and quantify myocardial iron distribution of FCM, a clinically available intravenous iron substitution agent, and to compare the myocardial distribution of FCM to that of the extracellular gadolinium-based contrast agent gadobutrol. We hypothesized that T1 mapping can be used to detect and quantify iron uptake in the myocardium.

## Material and methods

### Study population

The study was approved by the Stockholm Regional Board of Ethics (application number 2014/1140-31/3) and all study participants provided written informed consent. Healthy male volunteers 18 years of age or older were enrolled between October 2014 and October 2016. Healthy subjects were defined as having no previous history of cardiovascular or systemic disease and a normal resting electrocardiogram. Eight subjects underwent a CMR scan before and after intravenous administration of ferric carboxymaltose (Ferinject®, Vifor Pharma). This group was included in a prospective single-center study, using iron oxide as a contrast agent for cardiac magnetic resonance imaging (Iron-CMR) trial (Clinicaltrialsregister.eu registration no. EudraCT: 2014-002455-26). In addition, a separate group of seventeen healthy male subjects, who underwent CMR before and after intravenous administration of gadobutrol (Gadovist®, Bayer AB), were included, and this population has previously been reported as a part of a larger previous study [15]. Study exclusion criteria included a history of any chronic illnesses, cardiac diseases, presence of a pacemaker, claustrophobia, known allergy to either gadolinium-based contrast agent or hypersensitivity to iron products, liver dysfunction, or signs of iron overload.

### Image acquisition

CMR imaging was performed using a 1.5T scanner (Magnetom Aera, Siemens) with a 32-channel coil. In all subjects, T1 quantification was performed using a Modified Look-Locker Inversion-recovery (MOLLI) sequence. The protocol of a 5(3s)3 sampling scheme for pre-contrast imaging and a 4(1s)3(1s)2 sampling scheme [16] after intravenous administration of FCM was used, however the 5(3s)3 sampling scheme was used after intravenous administration of gadobutrol. T1 maps were reconstructed using inline motion correction [17]. The study population and the main component of CMR protocol used for calculation of FCM and gadobutrol lambda (λ) are outlined in Figure 1.Typical imaging parameters for MOLLI were matrix size 144×256 mm^2^, flip angle 35°, TE 2.6 ms, TR 278 ms, field of view 270×360 mm^2^, parallel acquisition technique (PAT) factor of 2, slice thickness 8 mm, phase encoding lines 125, FoV = 306×360 mm^2^.

**Figure 1.**
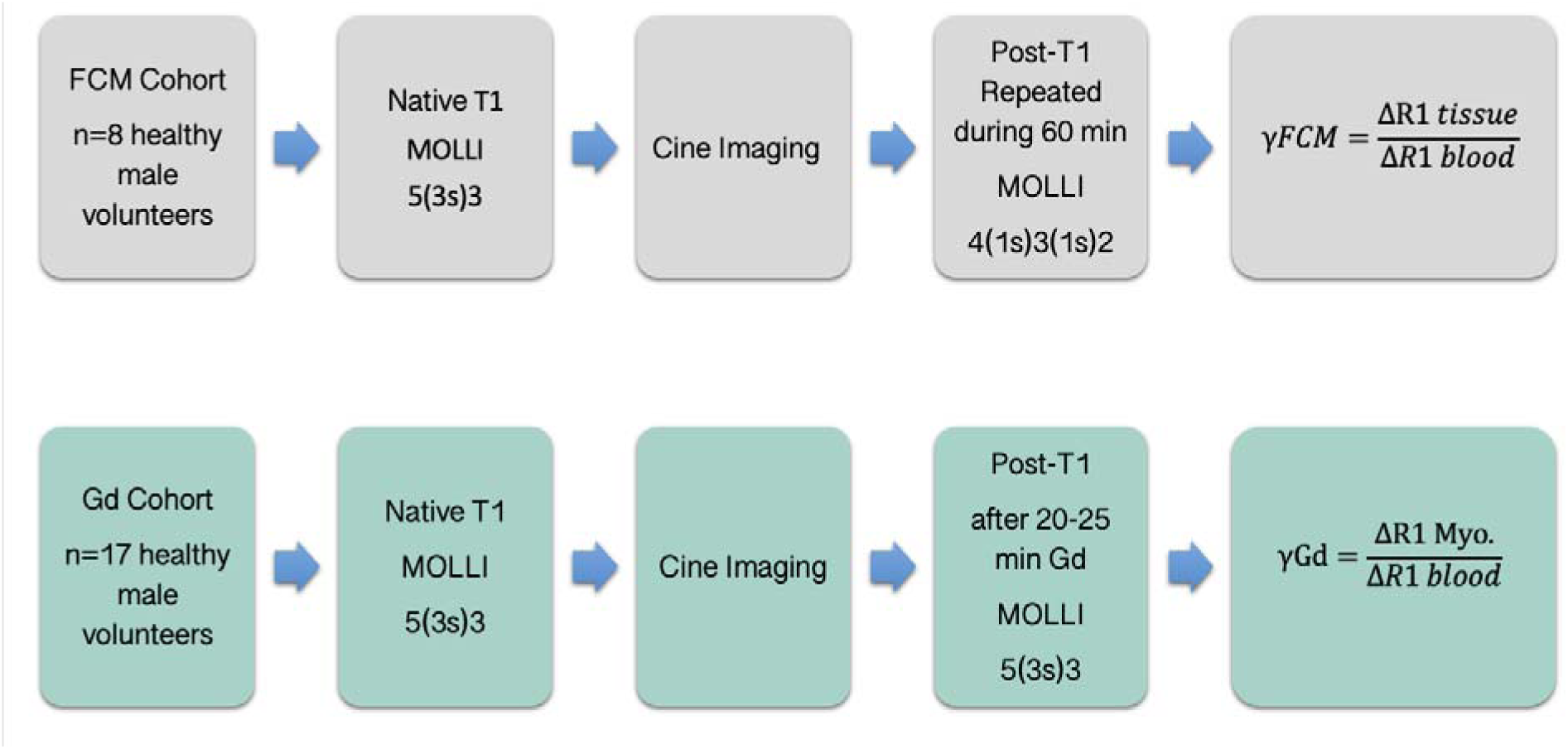
Study population and CMR protocol utilized for calculation of FCM and Gd lambda (λ). FCM, Ferric Carboxymaltose; Gd, gadolinium-based contrast agent, gadobutrol

T1 maps were acquired prior to the administration of the contrast agent, and then repeatedly during the 60-minutes period following the intravenous infusion of FCM. In this group, FCM (20 ml, 50 mg iron/ml, Vifor Pharma) was administered over a 15-minute-long infusion, followed by a bolus of saline (30 ml, 2 ml/s). In the gadolinium-based contrast agent group (n= 17), T1 maps were acquired before and around 15-20 minutes after intravenous administration of 0.2 mmol/kg of gadobutrol (Bayer AB) using the same T1 mapping imaging sequence as for the FCM group.

A balanced steady state free precession (bSSFP) cine sequence was acquired covering the left ventricle in a short-axis stack and three long-axis views, in order to calculate left ventricular volumes and function. Typical imaging parameters for cine imaging included: flip angle 57°, pixel size 1.4 mm^2^, TR/TE 2.4/1.2 ms, matrix size 143×256 mm^2^ and field of view 303 x 360 mm^2^.

### Image analysis

The quantification of left ventricular function, volumes and mass was performed offline using freely available software (Segment, Medviso AB) by two readers (GA and RT) with more than 5 years of experience in CMR imaging.

Furthermore, one reader (GA) measured T1 values in the myocardium, liver, spleen, skeletal muscle and the blood for the FCM group and T1 values of the myocardium and the blood pool for the gadobutrol group. Myocardial T1 was measured in a midventricular short-axis image by manually drawing a region of interest (ROI) in the midmural region of the septum. Blood T1 was assessed by drawing a ROI in the LV cavity blood pool, with care taken to avoid inclusion of the papillary muscles. T1 of liver, spleen and skeletal muscle was measured by manually drawing a ROI in each of the tissues. Figure 2 shows an example of ROI placement in the different tissues. The partition coefficient lambda for a given contrast agent in the myocardium, liver, spleen or skeletal muscle was calculated as the ratio of ΔR1 of a given tissue over ΔR1 of the blood (λ = ΔR1_tissue_ /ΔR1_blood_), where R1=1/T1 and ΔR1 is the difference in R1 between post- and pre-contrast [14].

**Figure 2.**
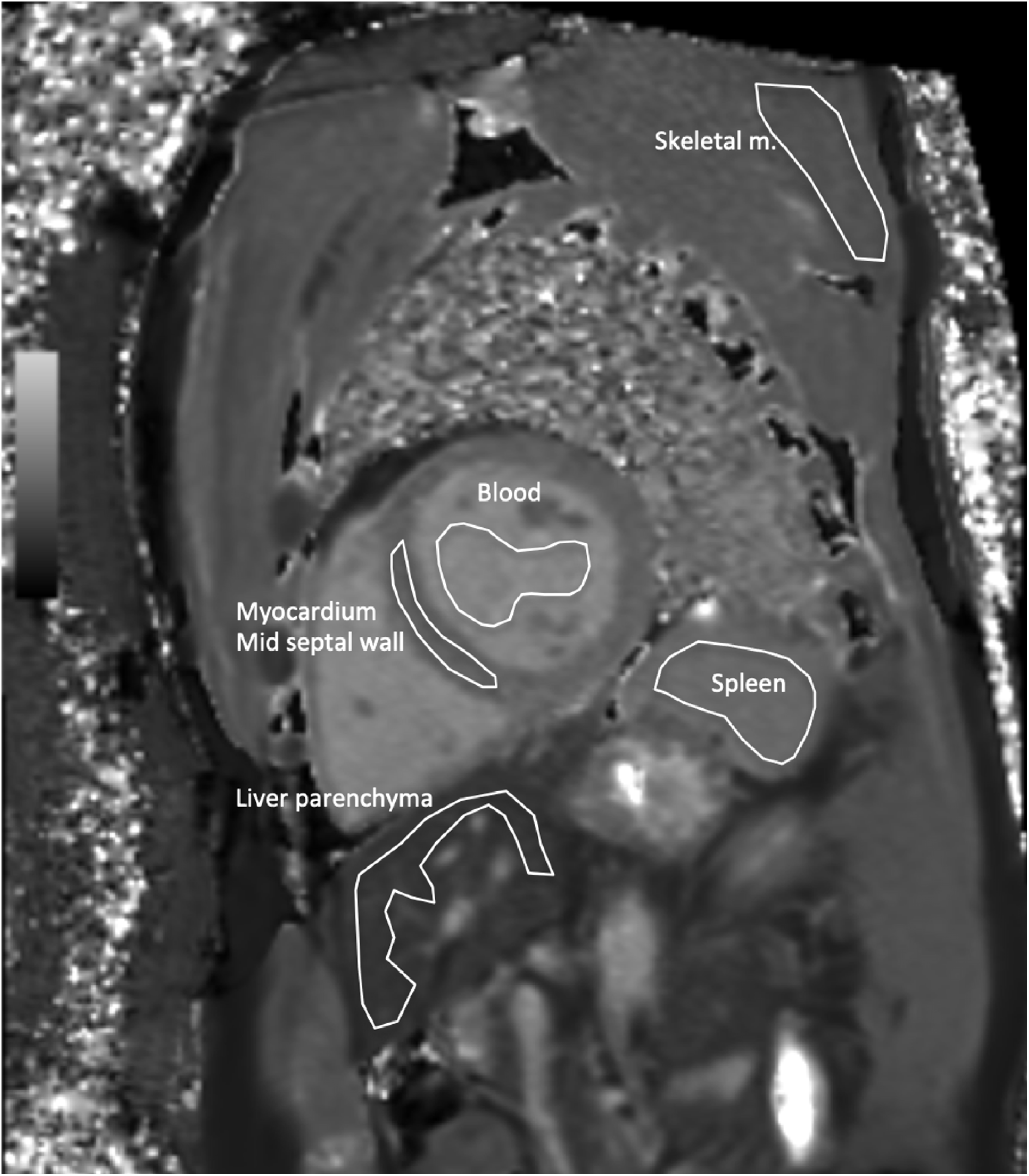
An example of regions of interest for T1 measurements made for each organ in a healthy volunteer. Note that the regions of interest for each organ were carefully placed to avoid partial volume.

### Statistical analysis

Statistical analysis was performed using SPSS^®^ software (version 24; IBM^®^, Somers, New York, USA) and Microsoft Excel^®^ (Microsoft^®^, Redmond, Washington, USA). Continuous data is presented as mean ± standard deviation (SD) or mean ± standard error of the mean (SEM) as designated. T1 measurement changes over time were tested using ANOVA. Differences between groups was tested using the two-tailed paired or unpaired t-test, as appropriate. Statistical significance was defined as *p* < 0.05.

## Results

A total of twenty-five healthy male volunteers were evaluated, eight subjects (age 27 ± 3 years, range 22-32 years) were included in the FCM group and seventeen subjects (age 27± 5 years, range 20-39 years) in the gadobutrol group. Clinical characteristics of the two groups are summarized in Table 1. For the subjects in the FCM group, T1 values in all tissues were lower 20 minutes after the onset of intravenous administration of FCM as compared to pre-contrast T1 (blood 1331±73 vs 1558±60 ms, *p* < 0.001; myocardium 914±32 vs 978±30 ms, *p* < 0.001; liver 535±47 vs 550±42 ms, *p* < 0.001; spleen1005±71 vs 1095±66 ms, *p* < 0.001; skeletal muscle 807±22 vs 854±27 ms, *p* < 0.001). An example of native and post-contrast T1 maps following intravenous administration of FCM is shown in Figure 3. Consequently, at the 20-minute time point, R1 for all tissues was higher than R1 before contrast injection and therefore there was an increase in ΔR1 for all tissues (*p* < 0.001 for all). λ for the myocardium and spleen in the FCM group remained constant over time (*p* = 0.29 and *p* = 0.25 for trend, respectively), and the λ values 30 min after the onset of FCM administration were 64±8% and 81±20%, respectively. However, λ for the liver and skeletal muscle in the FCM group increased over time (*p* < 0.001 for trend for both tissues), indicating the absence of a dynamic equilibrium. Figure 4 illustrates the change over a 60-minute period in T1, R1, ΔR1 and λ for myocardium, liver, spleen and skeletal muscle after a 15 minutes long infusion of FCM.

**Figure 3.**
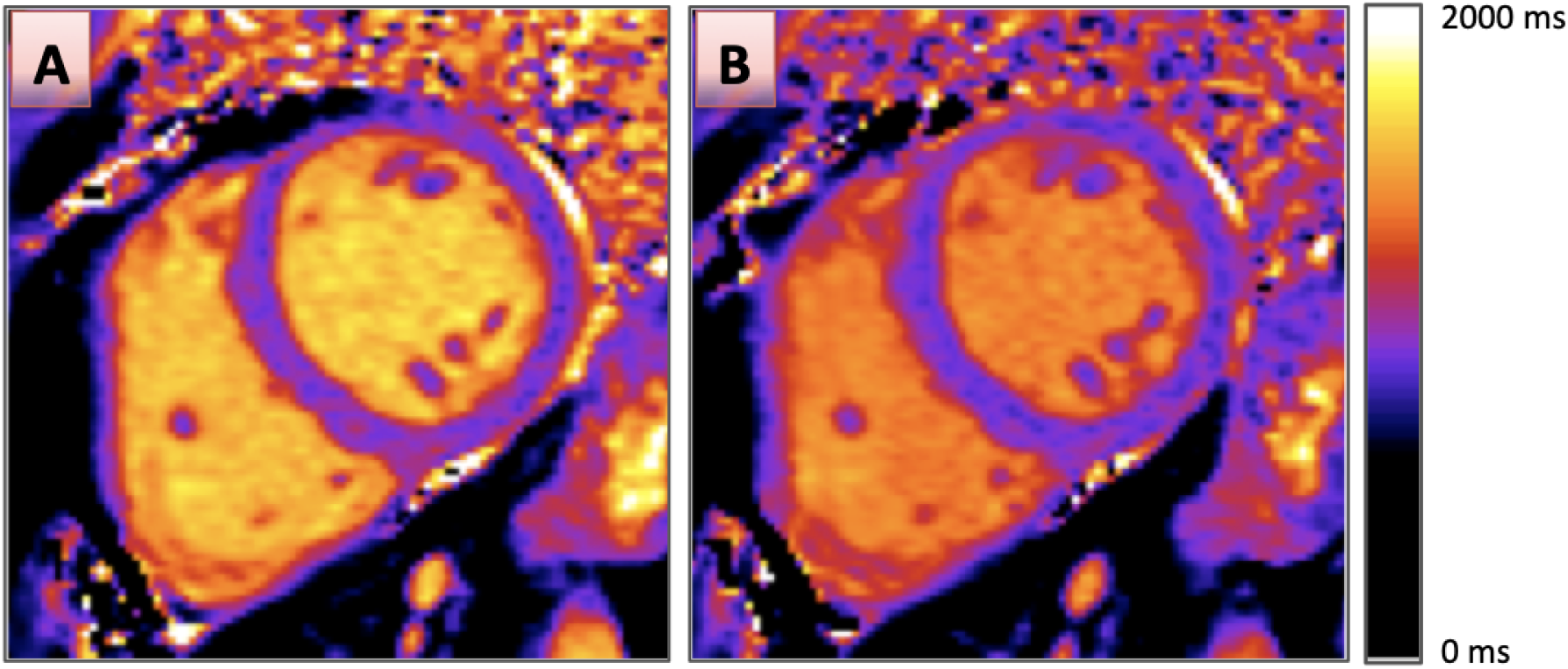
A pre-contrast native T1 map (A), and post-contrast (B) (25 minutes after onset of intravenous injection of ferric carboxymaltose) short-axis T1 map in a mid-ventricular slice in a healthy volunteer. Both T1 maps are displayed using the same color scale as displayed on the right. Note how the post-contrast images have a visibly shorter T1, most prominent in the blood, but also present in the myocardium.

**Figure 4.**
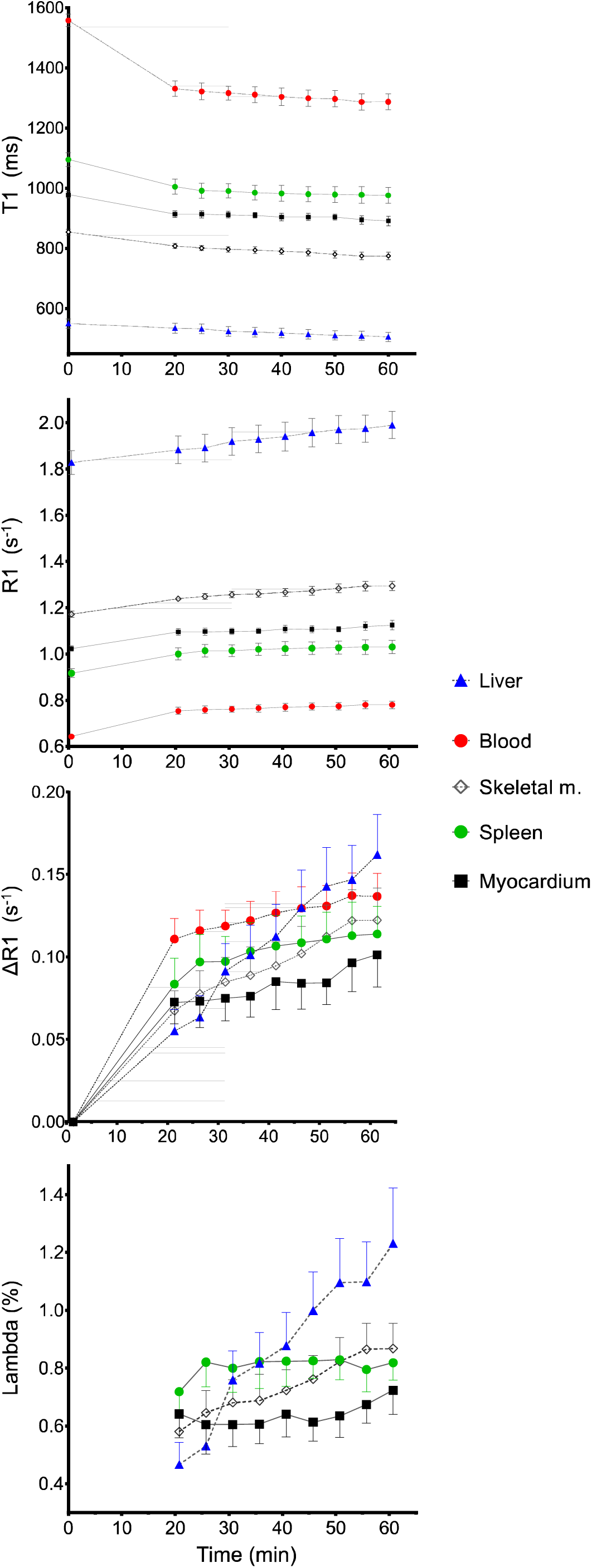
T1, R1, ΔR1 and the partition coefficient (lambda) of normal myocardium (black squares), liver (blue solid triangle), spleen (green solid circle), skeletal muscle (open diamond) and blood (red solid circles) over time after injection of ferric carboxymaltose in healthy volunteers (n=8). Note how lambda, which is the ratio of ΔR1 of a given tissue to ΔR1 of blood, remains constant over time for myocardium and the spleen, but not for the liver and skeletal muscle, up to 90 minutes after injection. Data are shown as mean values and error bars denote standard error of the mean.

**Table 1.** Subject characteristics.

|  | Ferric carboxymaltose | Gadobutrol | p-value |
| --- | --- | --- | --- |
| Male sex, n/total (%) | 8/8 (100) | 17/17 (100) | n/a |
| Age, years | 27±3 | 27±5 | 0.82 |
| Height, cm | 180±6 | 181±5 | 0.52 |
| Weight, kg | 81±13 | 79±9 | 0.59 |
| BMI, kg/m<sup>2</sup> | 25.0±3.5 | 24.0±2.7 | 0.41 |
| BSA, m<sup>2</sup> | 2.0±0.2 | 2.0±0.1 | 0.76 |
| LV end-diastolic volume, ml | 204±29 | 202±35 | 0.89 |
| LV end-diastolic volume index, ml/m<sup>2</sup> | 101±13 | 101±15 | 0.97 |
| LV end-systolic volume, ml | 77±12 | 82±18 | 0.44 |
| LV end-systolic volume index, ml/m<sup>2</sup> | 38±5 | 41±8 | 0.36 |
| LV ejection fraction, % | 62±3 | 59±4 | 0.08 |
| LV mass, g | 127±25 | 141±22 | 0.16 |
| LV mass index, g/m<sup>2</sup> | 64±10 | 70±9 | 0.15 |

Data are shown as mean±SD or number (%). BMI, body mass index; BSA, body surface area; LV, Left ventricle.

For the gadobutrol group, the λ of the myocardium measured at between 10-15 minutes after intravenous injection was 45±1%. Lambda of myocardium in the FCM group was higher than the λ of myocardium in the gadobutrol group (64±8 vs 45±1%, *p* = 0.003) (Figure 5).

**Figure 5.**
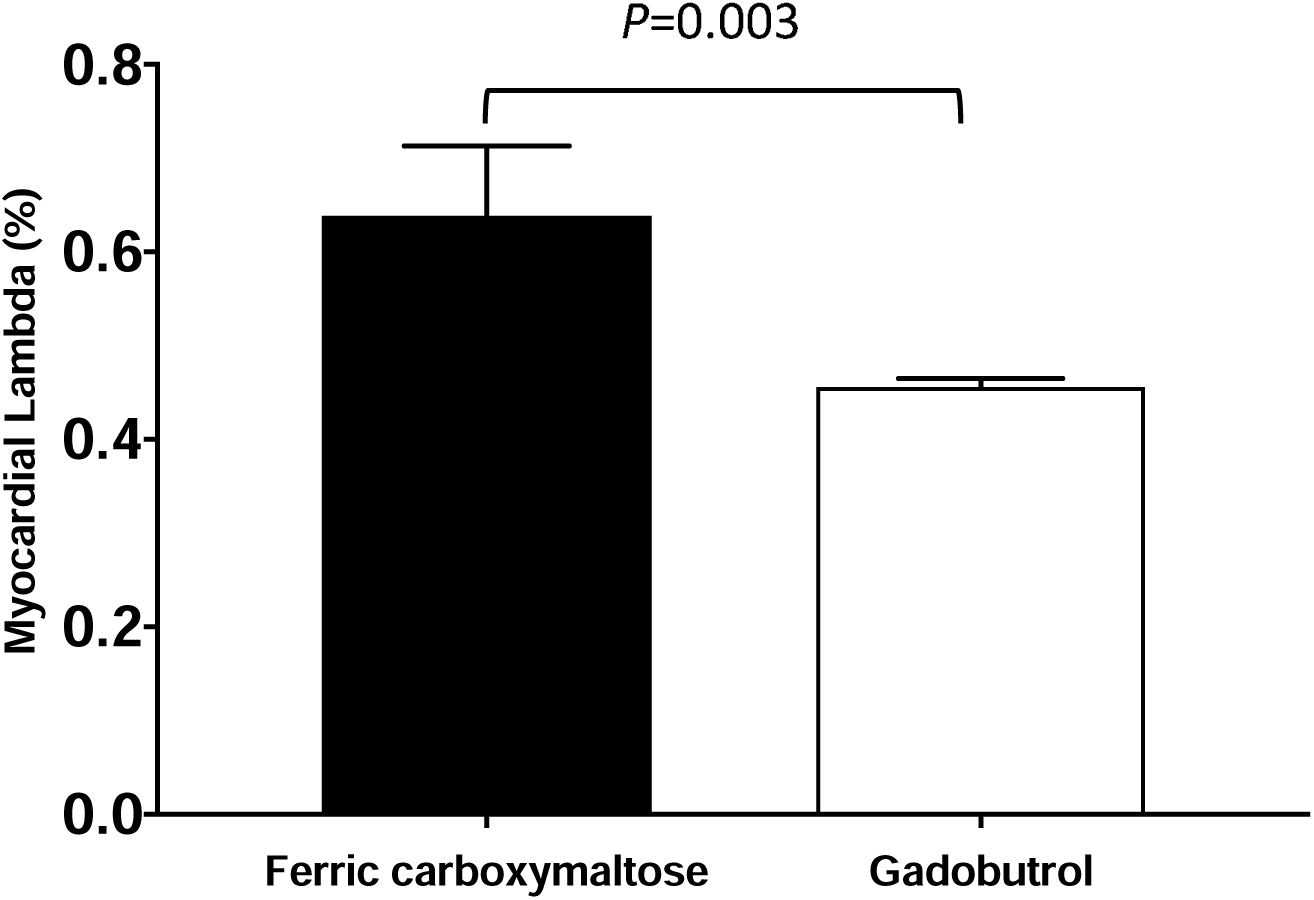
The fractional distribution volume (lambda) of ferric carboxymaltose and gadobutrol in healthy myocardium. Lambda in healthy myocardium for FCM was markedly higher than lambda for the extracellular agent, gadobutrol (P=0.003), illustrating a higher myocardial distribution of FCM compared to the extracellular distribution of gadobutrol. Error bars denote SEM.

## Discussion

The main finding of the study is that T1 mapping can quantitatively measure the *in vivo* distribution of intravenously administered FCM into the myocardium, liver, spleen, skeletal muscle and blood. Furthermore, the distribution of FCM into the myocardium was markedly higher than for the extracellular gadobutrol, suggesting an intracellular distribution for FCM.

R1 increases in all measured tissues following intravenous administration of FCM. This is consistent with a previous animal study using iron dextran loading that showed a linear relationship between R1 and tissue iron concentration in the liver and heart [18]. FCM administration is known to initially lead to systemic release of iron within the cells of reticuloendothelial system into the blood [19; 20]. The increased iron concentration in the blood following FCM administration is in agreement with a previously published human study [21]. Similarly, the current study shows that iron concentrations increase at different rates among the measured tissues.

A dynamic equilibrium [22], also referred to as an equilibrium distribution [13], is defined as an unchanged λ between tissue and blood over time after intravenous administration. This implies that the exchange of contrast agent between the blood and the tissue is more rapid than change of contrast agent concentration in the blood pool. The current study found that λ values in the myocardium and spleen did not change over time after intravenous administration of FCM (64±8% and 81±20%, respectively). This indicates that there is a dynamic equilibrium between the concentration of FCM in the myocardium, spleen and the blood. However, in the liver and skeletal muscle, such a dynamic equilibrium state was not achieved, and λ increased successively over time. These findings indicate that there is a continuously increasing concentration of FCM in the liver and skeletal muscle. This is likely due to the mechanisms and time course of physiological uptake of iron in the liver and skeletal muscle. Under physiological condition, non-heme iron is transported into cells by two main routes, namely, through transferrin-bound iron uptake [23] and non-transferrin-bound uptake such as through the divalent metal transporter 1 [24]. It may be that these mechanisms have a more drawn-out time course in the liver and skeletal muscle such that a dynamic equilibrium is not achieved during the first hour after intravenous administration. By comparison, in the setting of myocardial iron overload, the calcium channels have been suggested to be the main route of iron uptake into cardiomyocytes [25; 26]. Regardless of the fact that the exact mechanism may not be known, the data in the current study indicate that the time course of distribution in the myocardium and spleen is rapid enough to achieve a dynamic equilibrium. The current study has demonstrated that λ for FCM in healthy myocardium was on average 19 percentage points higher than λ for gadobutrol. This indicates that FCM distributes to a greater extent into the myocardium than extracellular agents like gadobutrol. Since extracellular agents already distribute exclusively into the extracellular space, the only additional compartment to which FCM can distribute into beyond that is the intracellular space.

In light of a relatively new recommendation of iron therapy in heart failure patients [4], improved understanding of the iron physiology that contributes to improvement of cardiac function in these patients’ group is of increasing importance. Quantification of myocardial λ of FCM by using T1 mapping may provide additional insight into the pathophysiology of heart failure and may provide more information on how the FCM influences myocardial function. However, further studies are required to evaluate the utility of T1 mapping for quantifying FCM in myocardium, specifically in patients with heart failure.

In terms of limitations, we did not compare the T1 values with simultaneously measured T2* which has been shown to have an agreement with tissue iron content [27], and is more specific for iron using single time point measurements. However, a recent study has shown that T1 mapping may be more sensitive than T2* in quantification of myocardial iron overload [9] and has a better reproducibility [9; 28]. Based on previous myocardial T1 measurements [29], one can expect that the 5(3s)3 protocol used after intravenous administration of gadobutrol is slightly less accurate in T1 measurements compared to 4(1s)3(1s)2 protocol. However, this influence is likely negligible due to the calculation of the partition coefficient.

In conclusion, T1 mapping can detect and quantify the dynamic tissue distribution of iron from FCM in the myocardium, liver, spleen and skeletal muscle. Notably, lambda in healthy myocardium for FCM was markedly higher than lambda for gadobutrol, indicating that FCM distributes to a greater extent into healthy myocardium than extracellular contrast agents, most likely due to additional distribution into the intracellular space. The described technique opens a new possibility to use cardiac magnetic resonance to study *in vivo* myocardial iron distribution, both in healthy subjects and in subjects with myocardial disease.

## Data Availability

All data produced in the present work are contained in the manuscript

